# Same-day discharge versus overnight stay after pulmonary vein isolation: an assessment on clinical outcomes and healthcare utilization

**DOI:** 10.1101/2024.10.27.24316210

**Authors:** SR (Stacey) Slingerland, JLPM (Maarten) Van den Broek, DN (Daniela) Schulz, GJ (Gijs) van Steenbergen, LRC (Lukas) Dekker, AJ (Alexandre) Ouss, D (Dennis) van Veghel, the participating centres of the ablation registration committee of the Netherlands Heart Registration (NHR)

## Abstract

**Background:** Atrial fibrillation is increasingly prevalent and constitutes a severe economic and clinical burden. Pulmonary vein isolation (PVI) is an effective treatment. Evidence on the safety of same-day discharge (SDD) after PVI, instead of overnight stay (ONS), is limited.

**Methods & results:** This retrospective study uses data from PVI’s performed between June 2018 and December 2020 in the Netherlands. Baseline characteristics, clinical outcome data and health care utilization, extracted from two national databases, were compared between the implementation of an SDD protocol in a single centre and a national benchmark where majority is an ONS strategy. Descriptive and bivariate analyses were performed. We included data from 11,812 PVI’s; 1,360 in the SDD group and 10,452 for the ONS benchmark. The SDD protocol group performed 57.7% of PVI’s in SDD, the benchmark 5.3% (p<.001). The SDD protocol group performed more cryoballoon PVI (90.8% vs. 39.2%, p<.001). There were no differences in bleeding (p=.830), thromboembolic (p=.893) or vascular complications (p=.720), or cardiac tamponade (p=.634). Peri-procedural hospital stay was significantly shorter in the SDD protocol group (0.50 day vs. 1.52 days, p<.001), without a reallocation of healthcare to outpatient clinic (p=.230), emergency department (p=.132) or higher rate of readmission (p=.092).

**Conclusion:** The SDD protocol group with 57.7% SDD has similar complication rates and lower health care utilization, compared to the national ONS benchmark with 5.3% SDD, indicating that SDD is a safe and effective alternative for ONS in patients undergoing PVI. The 5.3% ONS in the benchmark suggests a potential reduction of nationwide healthcare utilization.

## Introduction

Atrial fibrillation (AF) is the most common cardiac arrhythmia, associated with high healthcare costs, morbidity and mortality (1,2). About two per cent of the general population suffers from AF and it is expected that this number will double or triple in the coming decades due to ageing and earlier detection (1,2). Catheter-based pulmonary vein isolation (PVI) is an effective first- or second-line treatment superior to antiarrhythmic drug therapy (3–7). PVI is the most commonly performed complex ablation procedure but is a resource-intense procedure and increasing in demand (8–10).

Due to advancements in PVI, procedural and clinical outcomes have seen impressive improvements and complications have decreased significantly. The average procedure time went down from up to six hours to slightly over one hour (11). Nevertheless, overnight stay (ONS) still is common practice in the majority of electrophysiology centres (12). The combination of the sheer amount of PVI procedures and costs of in-hospital stay and limited bed availability, has led the cardiology community to search for innovations to shorten the time spent in hospital. An infrastructural innovation deemed to improve the patient experience while decreasing costs is the development of same-day discharge (SDD) protocols (13–15). Although earlier research indicated the safety and feasibility of SDD, evidence is still limited to randomized controlled trial studies or small historical cohort studies (14–27). In, this study, using real-world observational data, we evaluate the clinical outcomes and healthcare utilization, during hospital admission and after discharge, following the implementation of an SDD strategy, in comparison to a national benchmark in which SDD is not commonly performed.

## Methods

### Study design

This retrospective study compares the implementation of an SDD protocol in a single centre (‘*SDD-protocol group*’) to data from a national benchmark where the majority has an ONS strategy (‘*ONS-benchmark’*) in the same period, using observational data from the Netherlands Heart Registration (NHR) and Dutch Hospital Data (DHD).

#### Inclusion and exclusion criteria

The SDD-protocol group includes patients undergoing both primary PVI and re-do PVI between June 2018 and December 2020. The ONS-benchmark comprises patients undergoing PVI (primary or re-do) from other electrophysiology centres in the Netherlands in the same period. Both groups include unique patients, counted once for either PVI or re-do PVI.

### Pulmonary vein isolation procedure in the SDD-protocol group

In the SDD-protocol group, all patients were admitted two hours prior to the procedure and placed on a *nil per os* for twelve hours. Cryoballoon (CB) ablation was the default technology for primary PVI; re-do procedures were performed with 3D mapping and radiofrequency (RF)-ablation. CB-ablation was performed through unilateral venous access, using two sheaths (8Fr and 12Fr). Re-do procedures required bilateral venous access (two 8Fr and one 6Fr) and a 2Fr arterial sheath for hemodynamic monitoring. Ultrasound guidance for venous access was not standard at the time. A specialized nurse monitored procedural analgesia and sedation. Both CB-PVI and re-do procedures with RF were performed under uninterrupted oral anticoagulation. With intravenous heparin, an active clotting time was maintained at >300s while dwelling in the left atrium. Protamine was administered after retracting the catheter to the right atrium, prior to the removal of the sheaths. Manual compression and a six-hour compression bandage ensured venous (and arterial) closure. A cardiac ultrasound was performed immediately after the procedure to assess pericardial effusion. The attending physician examined the groin before discharge and issued an ultrasound when deemed necessary.

### The adoption of the SDD strategy

The SDD-protocol group is from the Catharina Hospital in Eindhoven (CHE), the Netherlands. The SDD protocol was adopted in 2018. The PVI procedure was not changed with the introduction of the SDD protocol, with the exemption of the physical examination of the groin in the evening hours, typically 1 hour after removal of the pressure bandage, instead of the next day.

Procedures ending before 3PM and free from any (suspected) complications, lead to SDD by protocol; this group is called the SDD-group. In case of (suspected) complications or at the operator’s discretion, the discharge could be postponed, e.g. until potential complications were resolved or ruled out. This group is called the unexpected ONS-group. Patients whose procedure ended after 3PM, stayed overnight per protocol and were discharged <24h. A completion before 3PM was chosen to prevent discharge at night since the protocoled six-hour compression bandage and one-hour mobilization would otherwise lead to discharge after 10PM. This group is the expected ONS-group. The distinction between SDD, expected ONS and unexpected ONS is made using the SDD-protocol group’s hospital data. We are not aware of the (peri)procedural protocols of the hospitals in the ONS-benchmark.

### Endpoints per data source

#### Baseline characteristics and clinical outcomes

NHR is an independent nationwide clinical registry in which hospitals register a standard set of baseline, procedural and outcome data for all invasive cardiac procedures (28,29). On an annual basis, the NHR publishes outcome data of partaking hospitals to improve the quality, safety and transparency of care. The baseline characteristics from NHR are displayed in Table 1. The following outcome data, are used: 30-day mortality, bleeding complications during admission, thromboembolic complication <72h, vascular complication <30 days, cardiac tamponade <30 days and phrenic nerve injury (PNI) during admission. All definitions are aligned with ESC (30,31).

**Table 1:**
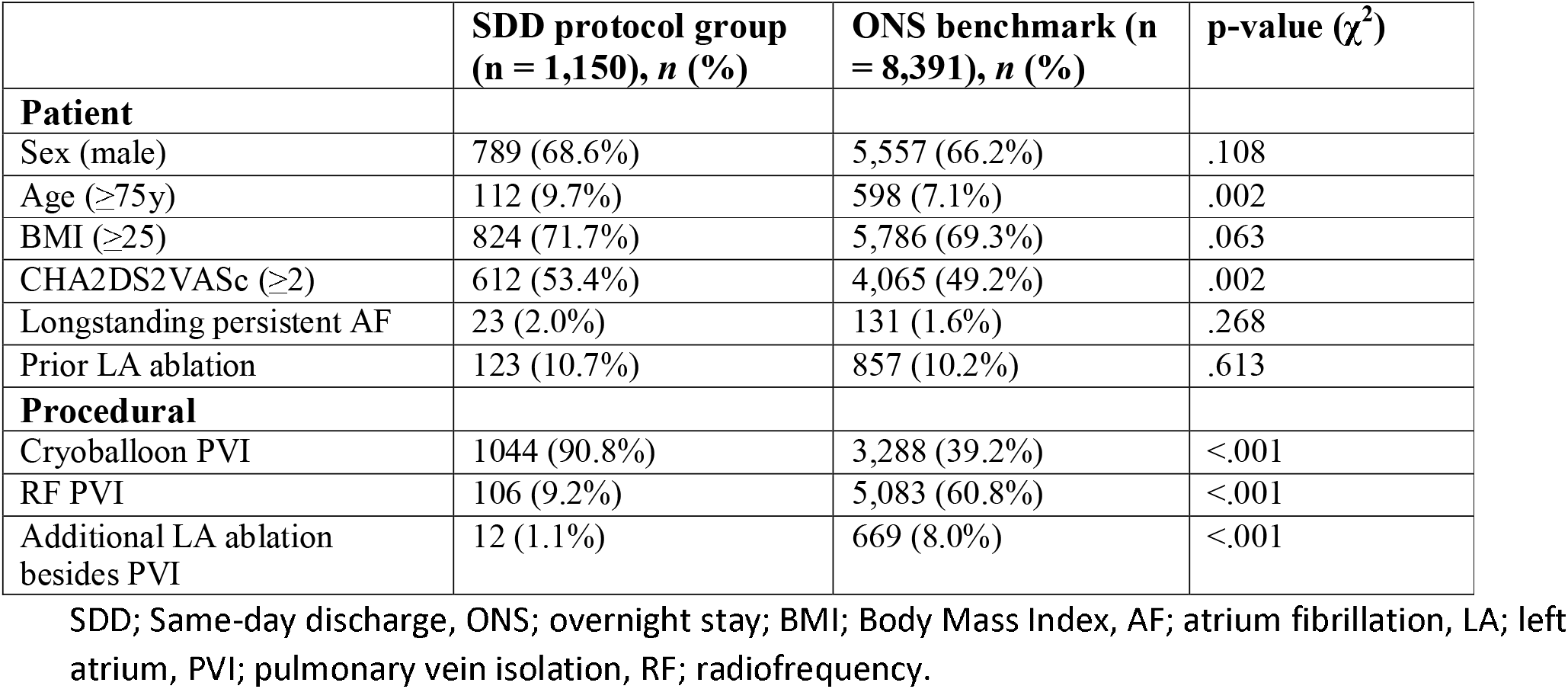
Baseline and procedural characteristics.

|  | **SDD protocol group**<br>**(n = 1,150), n (%)** | **ONS benchmark (n**<br>**= 8,391), n (%)** | **p-value ($\chi^2$)** |
| --- | --- | --- | --- |
| **Patient** |  |  |  |
| Sex (male) | 789 (68.6%) | 5,557 (66.2%) | .108 |
| Age ($\geq 75$y) | 112 (9.7%) | 598 (7.1%) | .002 |
| BMI ($\geq 25$) | 824 (71.7%) | 5,786 (69.3%) | .063 |
| CHA2DS2VASc ($\geq 2$) | 612 (53.4%) | 4,065 (49.2%) | .002 |
| Longstanding persistent AF | 23 (2.0%) | 131 (1.6%) | .268 |
| Prior LA ablation | 123 (10.7%) | 857 (10.2%) | .613 |
| **Procedural** |  |  |  |
| Cryoballoon PVI | 1044 (90.8%) | 3,288 (39.2%) | <.001 |
| RF PVI | 106 (9.2%) | 5,083 (60.8%) | <.001 |
| Additional LA ablation<br>besides PVI | 12 (1.1%) | 669 (8.0%) | <.001 |

SDD; Same-day discharge, ONS; overnight stay; BMI; Body Mass Index, AF; atrium fibrillation, LA; left atrium, PVI; pulmonary vein isolation, RF; radiofrequency.

#### Healthcare utilization

DHD manages medical and administrative data registered by Dutch hospitals to a national registry of hospital care (32). In this study, healthcare utilization is included as duration of peri-procedural hospital stay and healthcare utilization after discharge (emergency department (ED) and outpatient visits, in any Dutch hospital) to assess effects of relocation or delay of care after SDD implementation (with a follow-up of fourteen months after discharge).

### Statistical analyses

Patients were stratified to the SDD-protocol group or the ONS-benchmark. Statistical significant differences between the groups were assessed for categorical variables by a Chi-square test and a two-sided Student’s t-test for continuous variables. Results were considered statistically significant with a two-tailed p-value <α.05. All data analyses were performed in IBM SPSS V26.

## Results

According to the DHD data, 1,360 PVIs were performed between June 2018 and December 2020 in the SDD-protocol group and 10,452 in the ONS-benchmark. The SDD-protocol group performed significantly more procedures in SDD than the ONS-benchmark (respectively 57.7%, 5.3% p<.001). The results per year are shown in Table 1 and Figure 1 in the Appendix, showing the flowchart of the included patients over the different cohorts.

**Figure 1.**
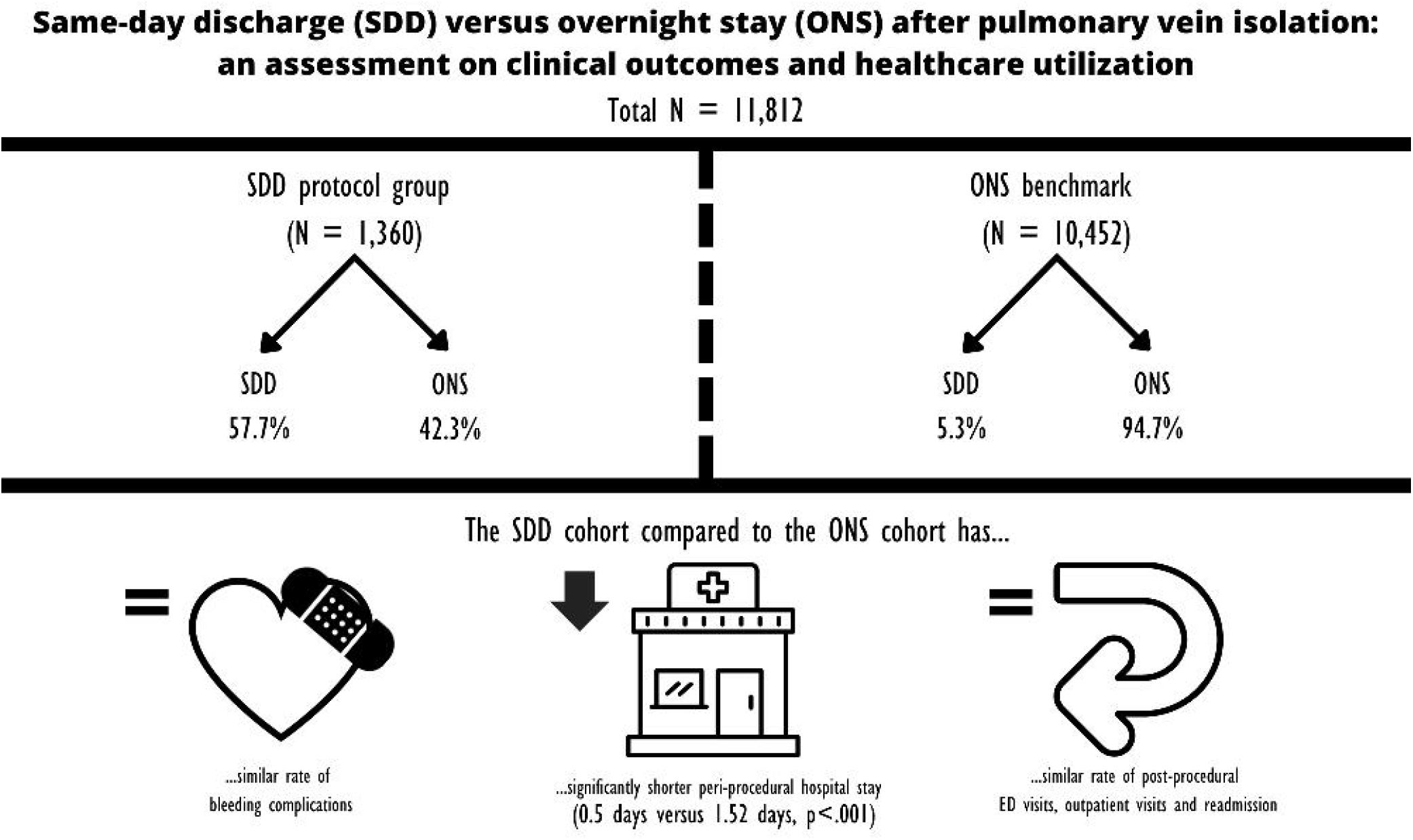

Table 1 shows the baseline and procedural characteristics of the SDD-protocol group (N = 1,150) and the ONS-benchmark cohort (N= 8,391), based on NHR data. Patients in the SDD-protocol group are significantly older and less healthy (higher CHA_2_DS_2_VASc-score) compared to the ONS-benchmark. Furthermore, the SDD-protocol group had more CB-PVI and fewer additional LA ablations, compared to the ONS-benchmark (p=.002, p=.002, p<.001, p<.001 respectively). Additional analyses on differences in baseline and procedural characteristics between SDD and (expected and unexpected) ONS cohorts within the SDD-protocol group are listed in Table 2 in the Appendix.

**Table 2:** Mortality, complications and healthcare utilization.

|  | SDD protocol group | ONS Benchmark | p-value |
| --- | --- | --- | --- |
| ***Clinical outcomes*** | ***N = 1,360*** | ***N = 10,452*** |  |
| 30-days mortality (n, %) | 0 (0.0) | 0 (0.0) | n/a |
| Bleeding complication during hospital stay (n, %) | 6 (0.5) | 42 (0.5) | .830 |
| Thromboembolic complication (<72h) (n, %) | 2 (0.2) | 17 (0.2) | .893 |
| Vascular complication (<30d) (n, %) | 14 (1.2) | 119 (1.5) | .720 |
| Cardiac tamponade (<30d) (n, %) | 5 (0.4) | 48 (0.6) | .634 |
| Phrenic nerve injury during admission (n, %) | 15 (1.3) | 49 (0.6) | .005 |
| ***Health care utilization*** | ***N = 1,150*** | ***N = 8,391*** |  |
| Peri-procedural hospital stay (days), (mean, SD) | 0.50 (1.13) | 1.52 (2.51) | <.001 |
| ED visit within 7 days after PVI (n, %) | 52 (3.82) | 495 (4.73) | .132 |
| Outpatient visits within 14 months, average visits (mean, SD) | 2.61 (2.34) | 2.74 (2.72) | .092 |
| Readmission within 4 months (n, %) | 214 (15.74) | 1,780 (17.03) | .230 |

SDD; same-day discharge, ONS; overnight stay, ED; emergency department

Table 2, using both DHD and NHR data, shows mortality, complications and healthcare utilization after PVI. There are no differences in mortality or complications except PNI; the ONS-benchmark has fewer events (0.6%) compared to the SSD protocol group (1.3%) (p = .005).

The mean duration of hospital admission was significantly shorter in the SDD-protocol group (0.50 days), compared to the benchmark (1.52 days)(p<.001). There is no statistical difference in healthcare utilization in terms of ED-visits <7 days after PVI, readmission <4 months or post-PVI outpatient visits <14 months.

## Discussion

This retrospective study, based on real-world data, compared clinical outcomes and healthcare utilization between a single centre SDD-protocol group and a national benchmark that relies mostly on an ONS strategy. Findings revealed no differences in mortality, bleeding complications or post-procedural healthcare utilization. Remarkably, these outcomes were consistent despite the SDD-protocol group comprising significantly older patients with a higher CHA2DS2VASc-score compared to the ONS benchmark. Furthermore, although peri-procedural hospital stay in the SDD-protocol group was significantly shorter than in the benchmark, this did not lead to a reallocation of care, evidenced by similar rates of hospital readmission, ED visits and outpatient visits. These results further contribute to the notion that SDD in this subpopulation is a safe and more efficient alternative to ONS with lower healthcare utilization.

The SDD-protocol group included a statistically significant higher proportion of CB-PVI procedures compared to the ONS-benchmark, 90.8% versus 39.2%, and hence had a statistically significantly higher percentage of PNI compared to the ONS-benchmark. PNI is a well-documented complication of CB-PVI, with previous studies indicating that CB-PVI is associated with up to ten times greater odds of PNI compared to RF-PVI (33–35). Importantly, PNI is often reversible and this study showed no differences in other clinical outcomes between the groups. This aligns with a recent meta-analysis by Huang et al., 2023, which demonstrated similar efficacy and safety between CB-PVI and RF-PVI (36). Moreover, despite differences in ablation methods, no difference in healthcare utilization were observed, indicating that PVI can be safely performed in SDD.

A common cause of concern when implementing SDD is the possible increase of complications occurring after discharge and/or the reallocation of healthcare utilization, such as more ED visits (37). However, our study, in accordance with recent studies, disputes this notion, showing no increase, substitution or relocation of healthcare utilization nor is there an increase in vascular complications (27,38).

Not all patients who were scheduled for SDD are discharged the same day, hence the unexpected ONS-group. Within the single centre that implemented a SDD protocol, the planned SDD was effectuated in 82.8% (Table 2, appendix), which is similar to results from other studies where planned SDD was effectuated in 79% to 99% of the cases (10,14–27). In the two studies with the highest percentage of effectuated planned SDD, recovery after PVI (including duration of groin compression bandage) was 3-4 hours as opposed to the 7 hours in the SDD-protocol group in this study (39,40). As expected, complications, such as PNI, occurred more often in the unexpected ONS-group, compared to the SDD-group. This complication is most often noted during the procedure, leading to a switch from planned SDD to unexpected ONS by protocol. Similarly, cardiac tamponade is another complication that is typically identified during the procedure and can also result in a switch from planned SDD to unexpected ONS. A slight 1% difference in cardiac tamponade is noted when comparing SDD versus ONS cohorts in general, which occurred more often in the ONS-group compared to the SDD group. This can also be attributed to the primary use of CB-PVI in the SDD group, as CB-PVI is associated with lower incidence of cardiac tamponade (41,42). In addition, the SDD-cohort contained significantly more female and younger patients with a lower CHA2DS2VASc-score and fewer prior LA ablation.

Lastly, SDD strategy was adopted to increase patient satisfaction, patient value and efficiency. The implementation of SDD will decrease healthcare costs since it prevents costs related to ONS such as usage of hospital beds during the night, night nurses and costs related to hospital admission (43). In times of scarce resources and rising costs, SDD after PVI is a safe and efficient way to provide healthcare for this group of patients, hence we recommend the implementation of SDD. The 94.7% of ONS in the benchmark indicates a potential reduction of nationwide healthcare utilization.

### Strengths, limitations and recommendations for future research

Several studies have demonstrated the safety, feasibility and cost-effectiveness of SDD-protocols (14–27). However, these studies often use historical cohorts, afternoon procedures or a single centre as the control group. This study’s strength lies in completeness and high-quality of the data from NHR and DHD, minimizing potential confounders such as the operator learning curve, and providing a larger control group compared to only using afternoon procedures. The national scope of the data ensures a representative view of the Dutch AF population undergoing PVI. The usage of DHD data allows complete follow-up information on all patients, including those from hospitals other than the one performing the PVI, reducing the risk of underestimating post-procedural events.

The differences in procedures and processes are an important limitation. The SDD-protocol group has a significantly higher percentage of CB-PVI compared to the benchmark, which could act as a confounder in this study. CB-PVI requires a larger diameter venous access sheath with a potential increased risk of access site complications (44,45). Except for PNI, RF and CB-PVI have been demonstrated to have an equal safety profile, hence we expect the effect of the energy source on post-discharge safety to be negligible (42). Furthermore, analysis of important procedural characteristics, such as protamine use, monitoring of activated clotting time and uni-or bi-lateral access, was not feasible, limiting our ability to correct for these potential confounders.

Another limitation is the number of inclusions from NHR is slightly lower than the number of DHD inclusions due to differences in registration; the NHR, utilizes ‘PVI’ as the inclusion criterion while in DHD the broader term ‘left atrium catheter ablation’ is used, thereby including e.g. left atrial tachycardia’s. However, these differences are minimal and their impact on the analyses is expected to be negligible. Second, DHD data were only provided at a group level, preventing correction for potential confounding factors. Nonetheless, bivariate analyses revealed minimal differences in patient and procedural characteristics, indicating the SDD-protocol group produced similar outcomes while treating an older population with higher stroke risk.

Another restraint of this study is that the extrapolation of our results might be limited. The SDD-protocol group is based in a high-volume, tertiary heart centre; the extensibility of these results towards low-volume and less experienced centres is uncertain and applying them to centres predominantly performing RF procedures is challenging.

For future research, it is recommended to add a costs-perspective to assess the total impact of the SDD implementation in terms of healthcare costs, especially in the context of rising healthcare costs, scarce resources and the need for efficient utilization.

## Conclusion

This study assessed differences in clinical outcomes and healthcare utilization following PVI after the implementation of an SDD strategy in a tertiary referral centre compared to a national benchmark that largely relies on an ONS strategy. Results demonstrate no difference in mortality and complications, except regarding PNI due to a higher percentage of CB-PVI. Peri-procedural hospital stay in the SDD-protocol group was significantly shorter than in the ONS-benchmark, without leading to a reallocation of healthcare to the ED department or outpatient clinic, resulting in a decrease of healthcare utilization. These results contribute to the notion that SDD in this subpopulation is a safe and effective alternative to ONS for patients undergoing PVI. Last, the percentage of ONS in the benchmark indicates a potential reduction of nationwide healthcare utilization.

## Data Availability

The data underlying this article were provided by the Netherlands Heart Registration (NHR) and Dutch Hospital Data (DHD) by permission. A request can be submitted to NHR and DHD to access these data.

## Author Contributions

SRS contributed to the paper by conceptualizing the study design, conducting formal analysis, interpreting results, and drafting the original manuscript. MB collaborated on data analysis, the analysis, interpretation of results, and drafted the manuscript.

During the literature review stage, DS, LD and GS provided valuable input. The manuscript underwent review and editing by DS, GS, LD and DV. LD, AO and DV provided project supervision. All authors have reviewed and approved the submitted version, and have agreed to be personally accountable for all aspects of the work in ensuring that questions related to the accuracy and integrity of any part of the manuscript are appropriately investigated and resolved.

## Acknowledgements

none

