## Supplementary Materials for "Same-day discharge versus overnight stay after pulmonary vein isolation: an assessment on clinical outcomes and healthcare utilization"

### Appendix Table 1

*Supplementary Table 1: Number of PVIs and average duration of hospital admission*

|  | **SDD protocol group (*n* = 1,360)** | | **ONS benchmark (*n* = 10,452)** | | **p-value (χ^2^ if applicable)** |
| --- | --- | --- | --- | --- | --- |
| **Year** | **SDD, *n* (%)** | **ONS, *n* (%)** | **SDD, *n* (%)** | **ONS, *n* (%)** |  |
| **Total** | 785 (57.7) | 575 (42.3) | 558 (5.3) | 9,894 (94,7) | <.001 (3726.37) |
| **2018** | 137 (42.6) | 185 (57.5) | 13 (0.5) | 2,519 (99.5) | <.001 (1013.58) |
| **2019** | 292 (60.6) | 190 (39.4) | 67 (1.6) | 4,186 (98.4) | <.001 (2013.45) |
| **2020** | 356 (64.0) | 200 (36.0) | 478 (13.0) | 3,189 (86.9) | <.001 (997.15) |
|  | Hospital stay (days), *mean (95% CI)* | | Hospital stay (days), *mean (95% CI)* | |  |
| **Total** | 0.50 | | 1.52 | | <.001 (14.7) |
| **2018** | 0.62 (0.52-0.72) | | 1.66 (1.52-1.79) | | <.001 |
| **2019** | 0.49 (0.36-0.62) | | 1.59 (1.52-1.67) | | <.001 |
| **2020** | 0.44 (0.36-0.51) | | 1.33 (1.28-1.38) | | <.001 |

### Appendix Table 2

*Supplementary Table 2:* *Patient characteristics and procedural and outcome variables comparing SDD and ONS within the SDD protocol group*

|  | **SDD (n = 696, 60.6%)** | **ONS (total)**  **(N=449, 39.4%)** | **p-value (χ^2^)** | **Unexpected* ONS (n = 145, 12.6%)** | **Expected* ONS (n = 304, 26.5%)** | **p-value (χ^2^)** |
| --- | --- | --- | --- | --- | --- | --- |
| **Patient N (%)** |  |  |  |  |  |  |
| Sex (male) | 494 (71.0) | 291 (64.8) | .029 (4.81) | 83 (57.2) | 208 (68.4) | .020 (5.38) |
| Age (≥75y) | 54 (7.8) | 58 (12.9) | .004 (8.23) | 19 (13.1) | 39 (12.8) | .935 (0.01) |
| BMI (>=25) | 496 (71.3) | 317 (70.6) | .809 (0.06) | 98 (67.6) | 219 (72.0) | .333 (0.94) |
| CHA2DS2VASc (>=2) | 350 (50.3) | 260 (57.9) | .012 (6.36) | 82 (56.5) | 178 (58.6) | .688 (0.16) |
| Longstanding persistent AF | 14 (2.0) | 8 (1.8) | .668 (0.18) | 3 (2.1) | 5 (1.6) | .751 (0.10) |
| Prior LA ablation | 59 (8.5) | 63 (14.0) | .003 (8.84) | 20 (13.8) | 43 (14.1) | .423 (0.63) |
| **Procedure, N (%)** |  |  |  |  |  |  |
| Cryoballoon PVI | 645 (92.7) | 395 (88.0) | .007 (7.24) | 125 (86.2) | 270 (88.8) | <.001 (241.59) |
| RF PVI | 51 (7.3) | 54 (12.0) |  | 20 (13.8) | 34 (11.2) |  |
| Additional LA ablation besides PVI | 5 (0.7) | 7 (1.6) | .173 (1.86) | 5 (3.4) | 2 (0.7) | .026 (4.98) |
| **Outcome, N (%)** |  |  |  |  |  |  |
| 30-day mortality | 0 (0) | 0 (0) | n/a | 0 (0) | 0 (0) | n/a |
| Bleeding complication during hospital stay | 3 (0.4) | 3 (0.67) | .587 (0.29) | 1 (0.7) | 2 (0.7) | .970 (0.003) |
| Thromboembolic complication (<72h) | 1 (0.1) | 1 (0.22) | .755 (0.10) | 0 (0) | 1 (0.3) | .490 (0.48) |
| Vascular complication (<30d) | 5 (0.7) | 9 (2.00) | .053 (3.74) | 3 (2.1) | 6 (1.9) | .965 (0.005) |
| Cardiac tamponade (<30d) | 0 (0) | 5 (1.11) | .005 (7.78) | 3 (2.1) | 2 (0.7) | .183 (1.78) |
| Phrenic nerve injury during admission | 9 (1.3) | 6 (1.3) | .950 (0.00) | 5 (3.4) | 1 (0.3) | .007 (7.25) |

**expected ONS; patients undergoing PVI who were primarily scheduled for ONS, unexpected ONS; patients undergoing PVI who were primarily scheduled for SDD.*

### Appendix Figure 1

*Supplementary Figure 1: Patient flow chart demonstrating the different SDD and ONS cohorts using both NHR and DHD data*

Study sample (N = 11,812)

Same Day Discharge (SDD) protocol group (N = 1,360) (a single electrophysiology center that implemented a SDD protocol)

Patients discharged in SDD: 785 (57.7%)

Patients discharged in ONS: 575 (42.3%)

Expected ONS (primarily scheduled for ONS): 67.7%

Unexpected ONS (primarily scheduled for SDD): 32.3%

Overnight Stay (ONS) national benchmark group (N = 10,452) (Dutch national benchmark of all other electrophysiology centres that follows an ONS strategy)

Patients discharged in SDD: 558 (5.3%)

Patients discharged in ONS: 9,894 (94.7%)

### Addendum

The following physicians are members of the Ablation Registration Committee of the NHR. They represent the hospitals that have provided the data for this study.

W. Kuijt, Amphia Ziekenhuis

A.H.G. Driessen, Amsterdam UMC

M. Kemme, Amsterdam UMC

P.H. van der Voort, Catharina Ziekenhuis

R.E. Bhagwandien, Erasmus MC

J. van der Heijden, HagaZiekenhuis

A. Adiyaman, Isala

A. Elvan, Isala

S.A.I.P. Trines, Leids Universitair Medisch Centrum

J.G.L.M. Luermans, Maastricht UMC+

K. Kraaier, Medisch Centrum Leeuwarden

P.F.H.M. van Dessel, Medisch Spectrum Twente

J.S.S.G. de Jong, Onze Lieve Vrouwe Gasthuis

S.W. Westra, Radboudumc

J.C. Balt, St. Antonius Ziekenhuis

R.J. Hassink, UMC Utrecht

Y. Blaauw, Universitair Medisch Centrum Groningen
